# High transferability of neutralizing antibodies against SARS-CoV-2 to umbilical cord blood in pregnant women with BNT162b2 XBB.1.5 vaccine - a retrospective cohort study

**DOI:** 10.1101/2024.03.18.24304517

**Authors:** Takuma Hayashi, Kenji Sano, Ikuo Konishi

**Author notes:** **Corresponding Address** Takuma Hayashi, Cancer Medicine, National Hospital Organization Kyoto Medical Center, 1-1, Fukakusa Mukaihatake, Fushimi-ku, Kyoto-city 612-8555, Kyoto.

## Abstract

Coronavirus disease 2019 (COVID-19), when contracted by pregnant women, can lead to severe respiratory illness, rapid disease progression, and higher rates of intensive care unit admission. COVID-19 infection during pregnancy is associated with an increased risk of preterm delivery, cesarean section, fetal dysfunction, preeclampsia, and perinatal death. Additionally, vertical transmission of severe acute respiratory syndrome coronavirus 2 (SARS-CoV-2) from pregnant women to their fetuses has been observed. While severe infections in neonates and infants are rare, newborns can experience serious consequences from COVID-19, despite their suboptimal humoral immune system protection. The amino acids in the structural proteins of SARS-CoV-2 are subjected to constant mutation. Since around January 2023, COVID-19, caused by infection with omicron-type SARS-CoV-2 variants, has been prevalent globally. Omicron-type SARS-CoV-2 variants can evade the immune response triggered by traditional mRNA-based COVID-19 vaccines, such as BNT162b2. Therefore, vaccination with a vaccine (BNT162b2 XBB.1.5) that can provide protection against omicron-type SARS-CoV-2 variants is recommended. Therefore, we examined the titers of anti-spike glycoprotein of SARS-CoV-2 IgG and IgA in the blood and umbilical cord blood obtained from pregnant women vaccinated with BNT162b2 XBB.1.5. The results showed that anti-spike glycoprotein of SARS-CoV-2 IgG and IgA titers were highest in the blood and cord blood obtained from pregnant women vaccinated with BNT162b2 XBB.1.5 at late gestational age (28–34 weeks). No serious side effects or adverse events caused by vaccination of pregnant women with BNT162b2 XBB.1.5 were observed in either pregnant women or newborns. In the future, to validate our findings, large cohort clinical studies involving numerous pregnant women must be conducted.

## Introduction

Severe acute respiratory syndrome coronavirus 2 (SARS-CoV-2) mutates repeatedly, and some new SARS-CoV-2 variants have spread rapidly and caused pandemics. In 2023, the omicron-type SARS-CoV-2 XBB.1.5 subvariant, an omicron subvariant created by the Ser486Pro mutation in the omicron-type SARS-CoV-2 XBB.1 subvariant, was spreading mainly in North America [1]. In the United States, the omicron-type SARS-CoV-2 BA.5 subvariant was the mainstream of coronavirus infections, which emerged in 2019 (COVID-19) until around November 2022, but from around January 2023, the BQ.1 subvariant and BQ.1.1 subvariant started to become prevalent. After that, the infection of the omicron-type SARS-CoV-2 XBB.1.5 subvariant spread rapidly [2].

Vaccination is the most effective prevention against many viral infections. However, each time a virus variant emerges, the preventive effects of the immune function and antiviral antibodies induced by vaccination are reduced [3]. A concerning issue with infections caused by SARS-CoV-2 is the aggravation caused by virus infection in elderly people and newborns, who might have low type 1 immune responses. Particularly, the transfer of anti-SARS-CoV-2 antibodies induced in the pregnant woman’s body by COVID-19 vaccination from the umbilical cord blood and/or breast milk to the fetus may be effective against neonatal COVID-19 [4,5].

COVID-19 vaccination (Pfizer/BioNTech; BNT162b2) during pregnancy produces functional anti-SARS-CoV-2 spike glycoprotein immunoglobulin G (IgG) and IgA in the circulation of pregnant women. These antibodies can also be detected in the umbilical cord blood at the time of birth of a newborn. Anti-SARS-CoV-2 spike glycoprotein IgG and IgA produced in the mother’s body through BNT162b2 vaccination during pregnancy may confer protection to newborns and infants against SARS-CoV-2 infection [6-9]. The titer of anti-SARS-CoV-2 spike glycoprotein IgG and IgA in cord blood correlates with the titer of these antibodies in maternal serum, with the highest levels reported after vaccination in the late second and early third trimesters [7-9]. XBB.1.5, one of the Omicron strain variants of SARS-CoV-2, is the most resistant virus variant to the immunity induced by BNT162b2 vaccination or SARS-CoV-2 infection. Consequently, Pfizer/BioNTech has received approval in many countries to manufacture and sell a vaccine (BNT162b2 XBB.1.5) specifically targeting the omicron-type SARS-CoV-2 XBB.1.5 subvariant. Therefore, COVID-19 vaccination (BNT162b2 XBB.1.5) tailored to the omicron-type SARS-CoV-2 XBB.1.5 subvariant is strongly recommended in each country. However, similar to BNT162b2 vaccination, serious side effects such as shock, anaphylaxis, myocarditis, and pericarditis have been associated with BNT162b2 XBB.1.5 vaccination. We compared and considered antibody titers of anti-SARS-CoV-2 spike glycoprotein IgG and IgA produced in the mother’s body by vaccination with BNT162b2 XBB.1.5, as well as antibody titers in umbilical cord blood of pregnant women after BNT162b2 XBB.1.5 vaccination.

## Result

Two doses of the omicron-type coronavirus vaccine (BNT162b2 XBB.1.5) administered to pregnant women may transfer antibodies to their babies. We measured the levels of neutralizing antibodies against SARS-CoV-2 in postnatal blood and cord blood obtained from the umbilical cords of 148 pregnant women who had received two doses of the BNT162b2 XBB.1.5 vaccine. Our results revealed that the level of neutralizing antibodies against SARS-CoV-2 in the umbilical cord blood (IgG Median 3,115.8 U/mL, IgA Median 2,234.5 U/mL) was 1.696 times and 1.353 times higher, respectively, than that in the mother’s blood (IgG Median 1,837.3 AU/mL, IgA Median 1,651.0 AU/mL) (Figure 1, eTable 1. Supplementary). Additionally, findings from a clinical study conducted in the United States supported these observations, showing that the level of neutralizing antibodies against SARS-CoV-2 in the umbilical cord blood (IgG Median 2,170 AU/mL) was 1.05 times higher than that in the mother’s blood (IgG Median 2,070 AU/mL).

**Figure 1.**
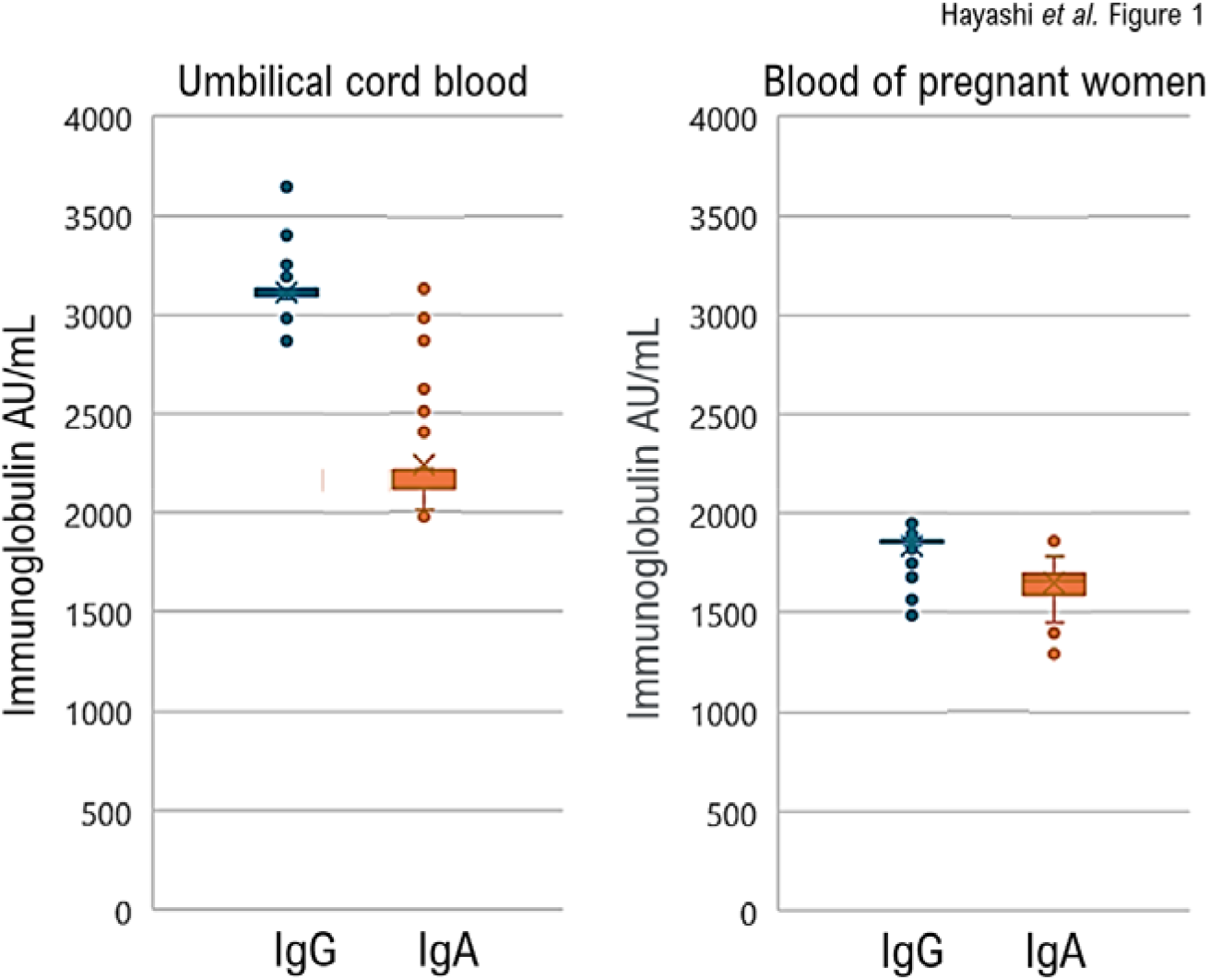
Changes in Levels of anti-spike glycoprotein of SARS-CoV-2 immunoglobulin A and immunoglobulin G antibodies in blood of pregnant women and umbilical cord blood during childbirth. Comparison of anti-SARS-CoV-2 IgA and IgG titers in blood and umbilical cord blood obtained from pregnant women at the time of delivery revealed that the anti-SARS-CoV-2 IgA and IgG titers in umbilical cord blood were significantly higher than those in maternal blood.

In a clinical study conducted by our facility in Japan, although the cohort was small, there is a possibility that compared to the amount of neutralizing antibodies against SARS-CoV-2 in the cord blood of American pregnant women who received two doses of the BNT162b2 XBB.1.5 vaccine, the neutralizing antibodies against SARS-CoV-2 produced in the bodies of Japanese pregnant women who received the same vaccine are more readily transferred to the umbilical cord blood. This finding suggests that anti-SARS-CoV-2 neutralizing antibodies produced in the bodies of pregnant women vaccinated with the BNT162b2 XBB.1.5 vaccine may be transferred to the fetus through the placenta.

Additionally, we investigated the optimal timing during pregnancy for administering the second dose of the BNT162b2 XBB.1.5 vaccine to pregnant women, aiming to achieve the highest levels of neutralizing antibodies against SARS-CoV-2 transferrable from the placenta. We found that the period of pregnancy with the highest levels of neutralizing antibodies against SARS-CoV-2 (IgG Median 3,320 AU/mL, IgA Median 2,632.5 AU/mL) was between 28 and 34 weeks of pregnancy (Figure 2, eTable 1 in Supplementary). Nonetheless, it was evident that even in pregnant women who received the BNT162b2 XBB.1.5 vaccine during early pregnancy or close to childbirth, a relatively high level of neutralizing antibodies against SARS-CoV-2 was transferred to the fetus through the placenta.

**Figure 2.**
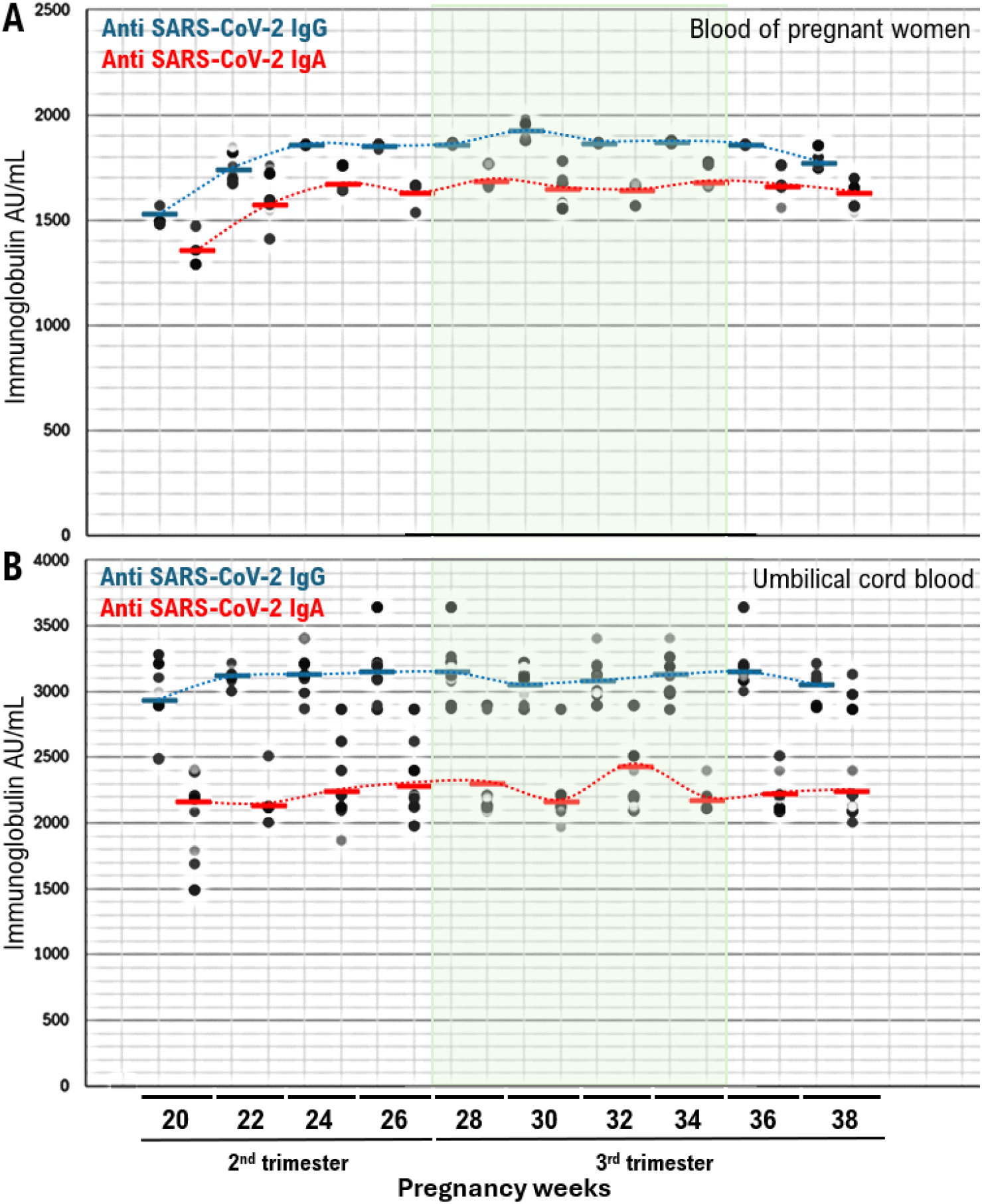
Anti-spike glycoprotein IgA and IgG titers in blood and umbilical cord blood obtained from pregnant women vaccinated with the BNT162b2 XBB.1.5 vaccine, shown by gestational age at the time of vaccination. **A**. Correlation between the time interval from the second COVID-19 vaccine dose and anti-SARS-CoV-2 immunoglobulin A and immunoglobulin G antibody levels in the blood of pregnant women. **B**. Correlation between the time interval from the second COVID-19 vaccine dose and anti-SARS-CoV-2 immunoglobulin A and immunoglobulin G antibody levels in umbilical cord blood.

Demographic and clinical characteristics are presented in Table 1. We defined severe neonatal morbidity (SNM) using an adaptation of the validated composite Neonatal Adverse Outcome Indicator, which includes 15 diagnoses and 7 procedures during the birth admission or within the first 28 days after birth. The risks of SNM, neonatal death, and Neonatal Intensive Care Unit admission were lower in infants of mothers vaccinated with the BNT162b2 XBB.1.5 during pregnancy compared with unvaccinated mothers (eTable 2 in Supplementary); notably, after inverse probability of treatment weighting, significantly lower risks persisted. Systemic adverse effects following the first and second vaccine doses are indicated in eTable 3 in Supplementary. Systemic side effects postvaccination are common symptoms observed in women of the same age who are not pregnant.

**Table 1.**
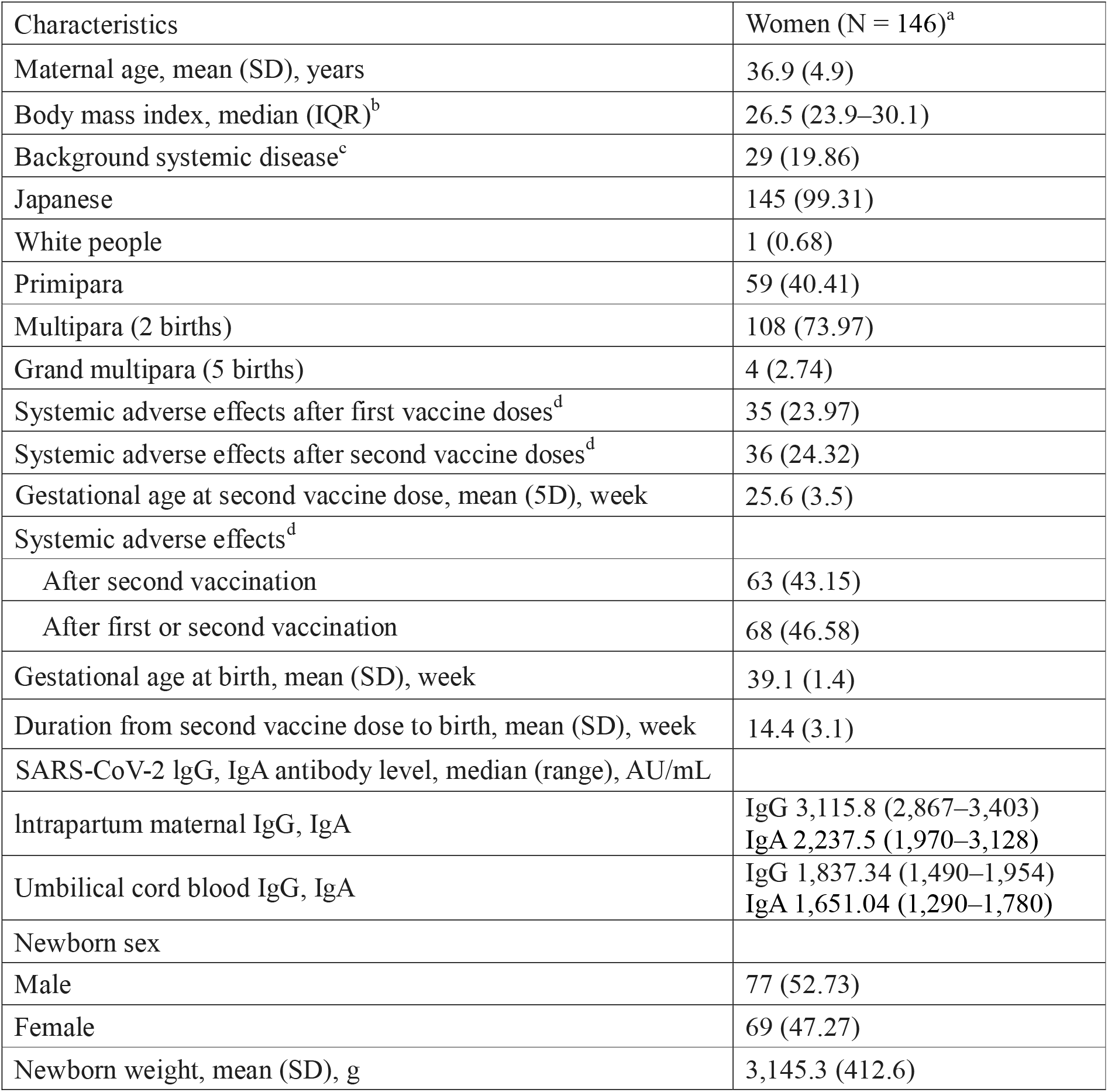
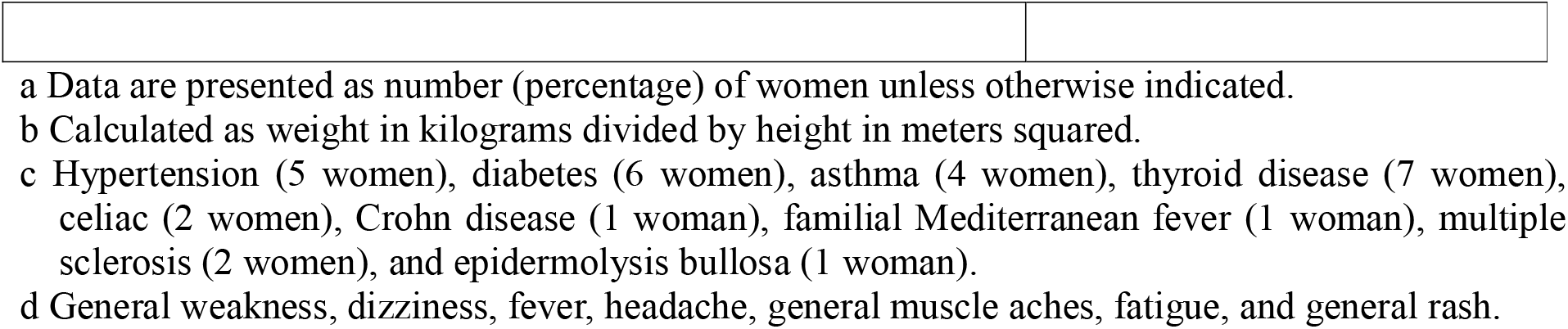
Demographic and Clinical Characteristics of Pregnant Women Vaccinated with the BNT162b2 XBB.1.5 mRNA COVID-19 Vaccine

| Characteristics | Women (N = 146)<sup>a</sup> |
| --- | --- |
| Maternal age, mean (SD), years | 36.9 (4.9) |
| Body mass index, median (IQR)<sup>b</sup> | 26.5 (23.9–30.1) |
| Background systemic disease<sup>c</sup> | 29 (19.86) |
| Japanese | 145 (99.31) |
| White people | 1 (0.68) |
| Primipara | 59 (40.41) |
| Multipara (2 births) | 108 (73.97) |
| Grand multipara (5 births) | 4 (2.74) |
| Systemic adverse effects after first vaccine doses<sup>d</sup> | 35 (23.97) |
| Systemic adverse effects after second vaccine doses<sup>d</sup> | 36 (24.32) |
| Gestational age at second vaccine dose, mean (SD), week | 25.6 (3.5) |
| Systemic adverse effects<sup>d</sup> |  |
| After second vaccination | 63 (43.15) |
| After first or second vaccination | 68 (46.58) |
| Gestational age at birth, mean (SD), week | 39.1 (1.4) |
| Duration from second vaccine dose to birth, mean (SD), week | 14.4 (3.1) |
| SARS-CoV-2 IgG, IgA antibody level, median (range), AU/mL |  |
| Intrapartum maternal IgG, IgA | IgG 3,115.8 (2,867–3,403)<br>IgA 2,237.5 (1,970–3,128) |
| Umbilical cord blood IgG, IgA | IgG 1,837.34 (1,490–1,954)<br>IgA 1,651.04 (1,290–1,780) |
| Newborn sex |  |
| Male | 77 (52.73) |
| Female | 69 (47.27) |
| Newborn weight, mean (SD), g | 3,145.3 (412.6) |

When a pregnant woman receives the BNT162b2 XBB.1.5 vaccine during the third trimester of pregnancy, it has been observed that the titer of anti-SARS-CoV-2 antibodies in the blood and umbilical cord blood of pregnant women at the time of delivery were high. Notably, there was no correlation between gestational age, neonatal weight, gender, or systemic adverse effects following BNT162b2 XBB.1.5 vaccination, and the anti-SARS-CoV-2 antibody titers in the blood and umbilical cord blood of pregnant women at delivery (eTable 2 in Supplementary, eTable 3 in Supplementary). Therefore, Japan’s Ministry of Health, Labor, and Welfare, in alignment with the U.S. National Institute of Health, recommends BNT162b2 XBB.1.5 vaccination for pregnant women to prevent the severity of COVID-19 caused by SARS-CoV-2 infection in infants.

## Discussion

A clinical study revealed the presence of neutralizing antibodies against the spike glycoprotein of SARS-CoV-2 in umbilical cord blood and breast milk of pregnant women vaccinated with the mRNA-based COVID-19 vaccine (BNT162b2) (10,11,12,13,14,15). However, the amino acids in the spike glycoprotein of SARS-CoV-2 mutate rapidly (16), leading to the emergence of numerous mutant viruses and their subvariants of SARS-CoV-2. Among these variants, the omicron-type SRAS-CoV-2, which emerged in South Africa in November 2021, has demonstrated the ability to evade the immune response induced by vaccination with the mRNA-based COVID-19 vaccine (BNT162b2) (17,18). Consequently, in Japan, as in other countries worldwide, vaccination with the BNT162b2 XBB.1.5 vaccine, which is compatible with the omicron SARS-CoV-2 spike glycoprotein, is recommended (19). In line with these recommendations, we have similarly shown that pregnant women vaccinated with BNT162b2 XBB.1.5 exhibit the transfer of neutralizing antibodies (IgG and IgA) against the spike glycoprotein of SARS-CoV-2 to umbilical cord blood and breast milk (10,13,14,15).

A previous study reported that 96% of 27 pregnant women who received the COVID-19 vaccine tested positive for SARS-CoV-2 IgG antibodies at delivery (10). Additionally, infants born to women vaccinated at least 3 weeks before delivery also tested positive for SARS-CoV-2 IgG antibodies (10). Therefore, the positive correlation observed between the anti-SARS-CoV-2 spike glycoprotein IgG levels in maternal blood post BNT162b2 XBB.1.5 vaccination and those in umbilical cord blood supports previous findings. The efficacy of mRNA-based COVID-19 vaccination, such as BNT162b2 in reducing the severity of COVID-19 has been well-documented (16,17,18). Consequently, research into vaccinating pregnant women, particularly those at high risk of severe COVID-19, is paramount to mitigating its impact on maternal health. The purpose of COVID-19 vaccination in pregnant women is to minimize the severity of the disease in the mother and the fetus by ensuring adequate levels of anti-SARS-CoV-2 antibodies during pregnancy. Passive immunization through vaccination plays a crucial role in shielding the fetus from placental transmission of SARS-CoV-2 (20,21,22).

Our clinical study confirmed that the level of neutralizing antibodies against SARS-CoV-2 in umbilical cord blood (3,120 AU/mL) exceeded the level in the mother’s blood (1,860 AU/mL) by 1.68 times. Similarly, a study in the United States found that neutralizing antibody levels in umbilical cord blood (2,170 AU/mL) were 1.05 times higher than those in maternal blood (2,070 AU/mL) (23). Additionally, a study in Israel assessed neutralizing antibodies against SARS-CoV-2 in pregnant women vaccinated during the second trimester and their newborns. The antibody titer in the newborn was 2.6 times higher than that in the mother’s blood (10), indicating that COVID-19 vaccination during the second trimester is associated with elevated antibody levels in mothers and newborns. These findings highlighted the importance of early vaccination for maternal and fetal health (10). Differences in the anti-SARS-CoV-2 spike glycoprotein neutralizing antibody titers between maternal and umbilical cord blood in clinical studies across different countries may be attributed to variations in the gestational weeks and ages of pregnant women at the time of COVID-19 vaccine administration. Furthermore, studies have indicated that if a pregnant woman vaccinated against COVID-19 contracts SARS-CoV-2, the titer of anti-SARS-CoV-2 neutralizing antibodies in her blood may increase further (24).

In this clinical study, we measured the levels of anti-spike glycoprotein of SARS-CoV-2 antibodies (including IgG, IgA) in the blood samples of pregnant women and their umbilical cord blood. Vaccination with the BNT162b2 XBB.1.5 vaccine during the second half of pregnancy (28–34 weeks) resulted in the highest levels of anti-spike glycoprotein of SARS-CoV-2 antibodies in maternal blood. This immunological response was associated with the transfer of anti-SARS-CoV-2 neutralizing antibodies to cord blood after vaccination. These findings provide support for administering COVID-19 vaccination during the second trimester of pregnancy to ensure maternal protection and neonatal safety during the pandemic. However, the number of pregnant women who participated in our clinical trials was limited. Therefore, conducting larger-scale clinical studies with a more extensive cohort of pregnant women is necessary to draw definitive conclusions and further validate these findings.

## Data Availability

All data produced in the present work are contained in the manuscript

## Abbreviations

COVID-19: Corona Virus Infectious Disease, emerged in 2019
NIH: National Institution Health
IgG: immunoglobulin G
SARS-CoV-2: severe acute respiratory syndrome coronavirus-2

## Footnote

All authors are receiving medical ethics education. In addition, this study has been approved as a clinical medical study at each medical facility. The human serum samples used in this study were purchased from RayBiotech life (Peachtree Corners, GA, USA), and therefore Informed consent from the patient is required. When a patient participates in our clinical research and our medical staff collects blood, we must receive a consent form signed by the patient.

## Data Sharing

Data are available on various websites and have also been made publicly available (more information can be found in the first paragraph of the Results section).

## Ethics statement

This study was reviewed and approved by the Central Ethics Review Board of the National Hospital Organization of Japan (Meguro, Tokyo, Japan) and the Central Ethics Review Board of Kyoto University (Kyoto, Kyoto, Japan). The approved number for this study is 50-201504. In order to carry out this research, the authors attended a research ethics education course (e-APRIN) conducted by Association for the Promotion of Research Integrity (APRIN; Shinjuku, Tokyo, Japan). The approved numbers of e-APRIN are AP0000151756, AP0000151757, AP0000151758, AP0000151769.

## Disclosure

The authors declare no potential conflicts of interest. The funders had no role in study design, data collection and analysis, decision to publish, or preparation of the manuscript.

## Acknowledgments

We appreciate Dr. Zhang W (Roche Tissue Diagnostics, Tucson, AZ, USA.) for critical research assistance. We thank all medical staff for clinical research at Kyoto University School of Medicine and the National Hospital Organization Kyoto Medical Center. The authors would like to thank Enago (www.enago.com) for the English language review.

## Author Contributions

The following individuals have contributed to the content and elaboration of the STROBE statement: T.H. performed most of the experiments and coordinated the project. T.H. and K.S. conceived the study and wrote the manuscript. T.H. and I.K. provided with information on clinical medicine and oversaw the entire study.

## Funding

This clinical research was performed with research funding from the following: Japan Society for Promoting Science for TH (Grant No. 19K09840), START-program Japan Science and Technology Agency for TH (Grant No. STSC20001), and the National Hospital Organization Multicenter clinical study for TH (Grant No. 2019-Cancer in general-02), and The Japan Agency for Medical Research and Development (AMED) (Grant No. 22ym0126802j0001), Tokyo, Japan.

## Transparency Document

The transparency document associated with this article can be found in the online version at http://.

## Materials and Methods

### 1. Sample collection and Enzyme-linked immunosorbent assay (ELISA)

The study included individuals who received the BNT162b2 XBB.1.5 vaccine (Pfizer-BioNTech) targeting the omicron-type SARS-CoV-2 XBB.1.5 subvariant during pregnancy, between 20 to 32 weeks’ gestation. Participants were enrolled in a prospective study across 28 academic medical centers in Japan, with their infants subsequently included in this follow-up study conducted from September 20, 2023, to January 20, 2024. Individuals infected before vaccination were excluded from the study.

Matched maternal and umbilical cord serum samples were collected at birth. For infants of vaccinated mothers, capillary serum samples were obtained via a microneedle device at 2 months after birth, while samples for infants of mothers who were vaccinated and mothers who had been infected with SARS-CoV-2 were collected at 6 months postbirth. All participants received two doses of the BNT162b2 XBB.1.5 vaccine administered 21 days apart. Serum samples were collected before vaccine administration and then once weekly for 6 weeks, starting from week 2 after the first dose. These samples were kept frozen until analysis. IgG levels were detected using the Elecsys Anti–SARS-CoV-2 S serology assay and read on the Cobas e801 analyzer, with a level of more than 0.8 U/mL considered positive (La Roche Ltd). IgA levels were measured using the EUROIMMUN AG Anti-SARS-CoV-2 S Kit, and an extinction ratio of samples over calibrator of more than 0.8 was considered positive (eMethods in Supplementary materials).

Human serum samples collected from maternal blood and umbilical cord blood were used for the detection of SARS-CoV-2 antibodies, specifically IgA and IgG, using ELISA kits. For IgA analysis, the samples were initially diluted at a ratio of 1:25 in the appropriate sample dilution buffers. Subsequently, they were assessed using a semilJquantitative analysis ELISA kit following the manufacturers’ instructions (EUROIMMUN AG, Luebeck, Germany). In this assay, the microplate wells were coated with recombinant spike structural protein derived from the omicron-type SARS-CoV-2 XBB.1.5 subvariant. The results were interpreted by calculating a ratio of the extinction of samples over the extinction of the internal calibrator, with a ratio greater than 0.8 being deemed positive. For IgG analysis, we used the Roche Elecsys Anti-SARS-CoV-2 S quantitative serology assay on the Roche Cobas e801 Analyzer according to the manufacturers’ instructions (La Roche Ltd, Basel, Switzerland). A result exceeding 0.8 units/ml, based on internal calibration curves, was considered positive (eMethods in Supplement materials).

### 2. Clinical Research

We conducted a multi-center retrospective observational clinical study involving participants who underwent cancer genomic medicine at cancer medical facilities in Kyoto, Japan. This study was reviewed and approved by the Central Ethics Review Board of the National Hospital Organization Headquarters in Japan (Tokyo, Japan) on November 18, 2020 (Approval code NHO R4-04) and by the Kyoto University School of Medicine (Kyoto, Japan) on August 24, 2022 (Approval code M237).

### 3. Informed Consent

All participants provided their informed consent to participate in this study. Our clinical research adheres to the principles outlined in the Helsinki Statement.

### 4. Institutional Review Board Approval

#### Institutional Review Board Statement and Consent to Participate

The experiments involving human cancer genome information obtained from results by cancer genome gene panels were conducted at Kyoto University, affiliated hospitals, and the National Hospital Organization Kyoto Medical Center in accordance with institutional guidelines.

The study received approval from the Institutional Review Board (IRB) of Kyoto University (Approval code M192, H31-cancer-2) on April 05, 2014, and June 16, 2016, and the IRB of National Hospital Organization Headquarter (Approval code H31-cancer-2) on November 09, 2019, and June 17, 2022.

### 5. Ethical Compliance with Human

This manuscript contains personal and/or medical information about identifiable individuals. It also includes a case report/case history about an identifiable individual. All authors have confirmed that this manuscript is sufficiently anonymized according to our anonymization policy. Authors obtained consent from the participants. This study involves human participants and was approved by Ethics Committees and Institutional Boards. This study does not involve research with animals.

The authors attended research ethics education through the Education for Research Ethics and Integrity (APRIN e-learning program, eAPRIN). The completion numbers for the authors are AP0000151756, AP0000151757, AP0000151769, and AP000351128. Consent to participate was required as this research was considered clinical research.

### 6. Statistical Analysis

All data are presented as the mean with standard error of the mean. Normality was assessed using the Shapiro–Wilk test. For comparing two groups, either the unpaired two-tailed *t* test or the Mann–Whitney *U* test was used. Multiple comparisons were performed using either a one-way analysis of variance followed by a Tukey post hoc test or a Kruskal–Wallis analysis followed by a post hoc Steel–Dwass or Steel test. A *p*-value of less than 0.05 was considered statistically significant. All statistical analyses were conducted using JMP software (SAS Institute, Cary, NC, USA).

### 7. Data Availability

The data supporting the findings of this study are available from the corresponding author upon reasonable request.

Details of Materials and Methods are indicated in Supplementary files, which are available online.

## Notes

### Competing Interest Statement

The authors have declared no competing interest.

### Author Declarations

We conducted a multi-center retrospective observational clinical study involving participants who underwent cancer genomic medicine at cancer medical facilities in Kyoto, Japan. This study was reviewed and approved by the Central Ethics Review Board of the National Hospital Organization Headquarters in Japan (Tokyo, Japan) on November 18, 2020 (Approval code NHO R4-04) and by the Kyoto University School of Medicine (Kyoto, Japan) on August 24, 2022 (Approval code M237). All participants provided their informed consent to participate in this study. Our clinical research adheres to the principles outlined in the Helsinki Statement. Institutional Review Board Statement and Consent to Participate: The experiments involving human cancer genome information obtained from results by cancer genome gene panels were conducted at Kyoto University, affiliated hospitals, and the National Hospital Organization Kyoto Medical Center in accordance with institutional guidelines. The study received approval from the Institutional Review Board (IRB) of Kyoto University (Approval code M192, H31-cancer-2) on April 05, 2014, and June 16, 2016, and the IRB of National Hospital Organization Headquarter (Approval code H31-cancer-2) on November 09, 2019, and June 17, 2022.

