## supplementary data for "High transferability of neutralizing antibodies against SARS-CoV-2 to umbilical cord blood in pregnant women with BNT162b2 XBB.1.5 vaccine - a retrospective cohort study"

doi:

**eMethods.**

This supplemental material has been provided by the authors to give readers additional information about their work.

### eMethods

#### *Study design and participant selection*

Pregnant individuals were eligible for inclusion in the prospective longitudinal pregnancy cohort (KMC IRB #2019-001108a) if they were 20 or older; received the available COVID-19 mRNA vaccine (BNT162b2 XBB.1.5) during pregnancy; were able to provide informed consent; and delivered at either National Hospital Organization Kyoto Medical Center and 28 affiliated medical facilities participating in clinical research.

Individuals with a history of SARS-CoV-2 infection prior to vaccination were excluded. Matched maternal and umbilical cord sera were collected during the delivery hospitalization. Liveborn infants of enrolled pregnant participants were enrolled after birth (KMC IRB #2019-001108b).

In a cross-sectional study design, we sought to assess infant titers at 2 months or 6 months of life, as these are clinically relevant time points when pediatric vaccines may be given, and there may be theoretical concern about persistence of infant titer resulting in antibody interference with vaccine response. Infants born to pregnant individuals testing positive for SARS-CoV-2 between 20 and 32 weeks' gestation were enrolled as a comparator group for antibody transfer. Due to the timing of the ninth wave of the COVID-19 pandemic in Japan (late spring to early winter 2023) relative to the period of sample collection (early winter and early spring 2024), we could only capture the 4-month timepoint in infants born to mothers received the BNT162b2 XBB.1.5 vaccine (Pfizer-BioNTech).

#### ***Sample* collection *and processing***

Maternal and umbilical cord sera were collected via venipuncture (YourBio Health, Medford, MA) during delivery hospitalization. Serum separator tubes were centrifuged at 1000 g for 10 minutes at room temperature. Serum and plasma were aliquoted into cryogenic vials and stored at -80°C.

#### *Enzyme-linked immunosorbent assay (ELISA)*

Antibodies against the SARS-CoV-2 Spike glycoprotein were quantified using an ELISA. ELISA plates were coated with 500 ng/mL of Asp614Gly and Ser486Pro Spike glycoprotein (kindly provided by Erica Saphire) and incubated for 30 minutes at room temperature. Plates were washed with washing buffer (0.05% Tween-20, 400mM NaCl, 50mM Tris, pH 8.0) and blocked with a 0.1% Bovine Serum Albumin (BSA) solution for 30 minutes at room temperature. Plates were washed, and sample was added at a dilution of 1:100. Plates were incubated with sample at 37°C for 30 minutes. Plates were washed, and a horseradish peroxidase (HRP)-conjugated goat anti-human IgG antibody or HRP conjugated goat anti-human IgA antibody (Bethyl Laboratories, Montgomery, Texas, USA) was added for detection of Spike-specific IgG and Spike-specific IgA. Plates were incubated with secondary antibody for 30 minutes at room temperature and then washed. 3,3',5,5'-tetramethylbenzidine (TMB) was used to develop the ELISA () and sulfuric acid was used to stop the ELISA. Signal was read at 450 nm and Phosphate Buffered Saline (PBS)-background corrected from a reference wavelength of 570 nm. Detectable anti-Sike glycoprotein IgG or anti-Sike glycoprotein IgA was defined as any value greater than the sum of the mean value of assay negative controls (known SARS-CoV-2 negative, unvaccinated individuals) and 3 x the standard deviation of those samples. The ELISA kit used in this experiment is shown below. IgG levels were detected by the Elecsys Anti–SARS-CoV-2 S serology assay and read on the Cobas e801 analyzer with a level of more than 0.8 U/mL considered positive (La Roche Ltd) and IgA with the EUROIMMUN AG Anti-SARS-CoV-2 S Kit with an extinction ratio of samples over calibrator of more than 0.8 considered positive.

#### *Statistical Analyses*

Differences between vaccinated and naturally-infected groups depicted in Table 1 were assessed using Mann Whitney U test for continuous variables, or Fisher's exact test for categorical variables. $P<0.05$ was considered significant. Statistical analyses were performed using R, version 4.0.2, and Graphpad Prism v.9.0.

**eTable 2**. Association Between COVID-19 Vaccination: BNT162b2 XBB.1.5 During Pregnancy and Neonatal and Infant Outcomes

| | No./total No. (%) |
|---|---|
| Outcome | Vaccine exposed |
| Severe neonatal morbidity | 1/149* (0.67) |
| Neonatal death | 0/149 (0.00) |
| Neonatal intensive care unit (NICU) admission | 2/149 (1.34) |
| Neonatal readmission | 2/149 (1.34) |
| Discharged within 7 day after birth | 2/149 (1.34) |
| Hospital admission up to 6 month of age | 0/149 (0.00) |
| Discharged within 7 day after birth | 0/149 (0.00) |

*A pregnant woman who received the COVID-19 vaccine gave birth to twins.

**eTable 3.** 28-Day Risk of Adverse Events Following Vaccination With COVID-19 Vaccination: BNT162b2 XBB.1.5

| | BNT162b2 XBB.1.5 vaccine (risk period) |
|---|---|
| Outcome | No. of events/person-years |
| Ischemic cardiac event | 1/148 (0.67%) |
| Any cerebrovascular event | 0/148 (0.00%) |
| Cerebral infarction only (including TIA) | 0/148 (0.00%) |
| Arterial thromboembolism | 0/148 (0.00%) |
| Deep vein thrombosis | 0/148 (0.00%) |
| Pulmonary embolism | 0/148 (0.00%) |
| Myocarditis | 0/148 (0.00%) |
| Pericarditis | 0/148 (0.00%) |
| Cerebral venous thrombosis | 0/148 (0.00%) |
| Thrombocytopenia and coagulative disorders | 1/148 (0.67%) |
| Guillain−Barre syndrome | 0/148 (0.00%) |
| Bell palsy | 0/148 (0.00%) |
| Transverse myelitis | 0/148 (0.00%) |
| Encephalomyelitis or encephalitis | 0/148 (0.00%) |
| Narcolepsy | 2/148 (1.35%) |
| Appendicitis | 0/148 (0.00%) |
| Aseptic arthritis | 1/148 (0.67%) |
| Type 1 diabetes | 0/148 (0.00%) |
| Subacute thyroiditis | 1/148 (0.67%) |
| Heart failure | 0/148 (0.00%) |
| Arrhythmia | 3/148 (2.03%) |
| Acute liver failure | 0/148 (0.00%) |
| Acute kidney failure | 0/148 (0.00%) |
| Acute pancreatitis | 0/148 (0.00%) |
| Erythema multiforme | 1/148 (0.67%) |
| Seizure | 2/148 (1.35%) |
| Arterial aneurysm | 0/148 (0.00%) |
| Uveitis | 2/148 (0.67%) |
| Muscle pain | 38/148 (25.68%) |
| Arthro joint pain | 34/148 (22.97%) |
| Fever | 22/148 (14.86%) |
| Fatigue | 93/148 (62.84%) |
| Headache | 81/148 (54.73%) |
| Chills | 47/148 (31.76%) |
| Diarrhea | 3/148 (2,20%) |
